# Two-Year Evolution of a Prospective Audit and Feedback Antimicrobial Stewardship Program in a Quaternary Intensive Care Unit in Ghana

**DOI:** 10.64898/2026.07.11.26357812

**Authors:** Peter Kwamina McCarthy, Nana Ama Buadiba Osei, Daniel Freeman Owusu Ansah, Josephine Mensah, Salomey Asaah Denkyira, Fareeda Serwaa Brobbey, Glennsa Nana Abena Ohene, Blessing Boakye Yiadom, George Boateng Kyei

**Affiliations:** University of Ghana Medical Centre, Accra, Ghana; Washington University in St. Louis, St. Louis, MO, USA 63130; Greater Accra Regional Hospital, Accra, Ghana

**Keywords:** antimicrobial stewardship, intensive care unit, prospective audit and feedback, clinician adherence, carbapenem use, Ghana, low- and middle-income countries

## Abstract

**Objectives:** To evaluate two-year, side-by-side outcomes of a prospective audit and feedback (PAF)-based antimicrobial stewardship program (ASP) in a quaternary ICU in Ghana, comparing diagnostic stewardship, antimicrobial prescribing patterns, and clinician adherence to stewardship recommendations between 2024 and 2025. Longitudinal PAF data from low- and middle-income countries (LMIC) quaternary ICUs are scarce; this study addresses that evidence gap.

**Methods:** A retrospective comparative analysis of routine Antimicrobial Stewardship (AMS) surveillance data was conducted at the University of Ghana Medical Centre ICU: 102 visits in 2024 and 63 in 2025. Proportions were compared by chi-square or Fisher’s exact test; continuous variables by Mann-Whitney U. Wilson score 95% confidence intervals (CIs) were computed for primary proportions.

**Results:** Biomarker-guided prescribing rose from 86.3% to 100% of visits (p=0.005) and culture and sensitivity testing from 74.5% to 90.5% (p=0.02). Targeted (culture-guided) therapy increased significantly from 23.5% to 41.7% of antibiotic recipients (p=0.03), while empiric prescribing declined correspondingly. Overall antibiotic utilization remained high in both years (96.1% vs 95.2%; p=1.00), and meropenem use rose from 42.9% to 56.7% (p=0.13). AMS interventions were recommended in 67.6% and 63.5% of visits, respectively. Clinician acceptance improved markedly from 40.6% (95% CI: 29.8–52.4%) to 67.5% (95% CI: 52.0–79.9%) (p=0.01).

**Conclusions:** Two years of PAF in a Ghanaian quaternary ICU demonstrated progressive program maturation: universal biomarker adoption, a significant shift toward targeted prescribing, and markedly enhanced clinician acceptance. Persistently high antibiotic utilization and rising carbapenem dependence underscore the need for sustained surveillance and carbapenem-sparing strategies in LMIC critical care.

## INTRODUCTION

Antimicrobial resistance (AMR) represents one of the most consequential threats to global public health, with intensive care units (ICUs) serving as high-stakes epicenters for the emergence and dissemination of MDROs.^1–3^ ICU patients frequently present with severe, polymicrobial, or culture-negative sepsis, necessitating empiric broad-spectrum antimicrobial therapy while awaiting microbiological confirmation. In the absence of structured oversight, this practice accelerates the development of resistance, increases rates of adverse drug events, prolongs hospital stays, and amplifies healthcare costs.^4^

The World Health Organization (WHO) and the Infectious Diseases Society of America (IDSA), in conjunction with the Society for Healthcare Epidemiology of America (SHEA), identify prospective audit and feedback as a cornerstone AMS strategy, complemented by formulary restriction, diagnostic stewardship, and multidisciplinary ward rounds.^5–8^ Prospective audit and feedback(PAF) is a core stewardship strategy that is proven to reduce the length of hospital stays and unnecessary antimicrobial use in hospitalized patients.^9–11^ When embedded in daily ICU rounding workflows, PAF has consistently reduced unnecessary antimicrobial days, improved de-escalation rates, and enhanced prescriber engagement.^12–14^ Yet despite this evidence base, longitudinal data tracking the real-world implementation and year-on-year evolution of PAF programs, particularly in quaternary ICU settings within low- and middle-income countries (LMICs), remain limited. Most published LMIC AMS data describe single-point-in-time surveys or short intervention periods, leaving critical gaps in understanding how PAF programs mature over time and which outcomes change most meaningfully.

Ghana, like many sub-Saharan African nations, faces compounding AMR pressures: high infectious disease burden, limited microbiological infrastructure, formulary constraints, and workforce shortages that complicate AMS implementation.^15^ The University of Ghana Medical Centre (UGMC)^16^ established a multidisciplinary AMS program in its ICU in 2024, providing a rare opportunity to track the program’s evolution using prospective surveillance data over two consecutive calendar years.

This study compares AMS surveillance outcomes, diagnostic utilization, antimicrobial prescribing patterns, and clinician adherence to stewardship interventions in the UGMC ICU over 2024 and 2025. Our objectives were to: 1-characterize the evolving patient and diagnostic profile of ICU AMS encounters; 2- quantify changes in antimicrobial prescribing patterns, with particular attention to carbapenem use; 3-measure the trajectory of clinician acceptance of AMS recommendations; and 4 - identify persistent prescribing challenges to inform future stewardship priorities.

## METHODS

### Study Design and Setting

A retrospective comparative analysis was conducted using routine AMS ward round surveillance data from the ICU at the University of Ghana Medical Centre, a quaternary referral and teaching hospital located in Accra, Ghana. The study period spanned January 1–December 31, 2024, and January 1–December 31, 2025. The data was assessed for research purposes between May 1, 2026, and May 30, 2026. The unit of analysis was the AMS ward round visit (hereafter “visit”), defined as a documented encounter in which the AMS team reviewed a patient during a scheduled prospective audit round. Patients reviewed on multiple occasions contributed multiple visits to the dataset, reflecting the longitudinal, ongoing nature of ICU stewardship surveillance. Authors had temporary access to identifiable patient information during data abstraction. Identifiers were removed before statistical analysis.

### Patient Population

All adult and pediatric patients reviewed during scheduled AMS ward rounds were included. Visits were identified through the AMS team’s daily prospective audit logs. There were 102 documented visits in 2024 and 63 in 2025. The planned visit-level analytical approach, rather than unique patient de-duplication, was pre-specified to capture the full scope of stewardship encounters and is consistent with the program’s primary operational objective of optimizing each prescribing decision at each clinical review.

### Data Collection

Standardized data extraction captured: patient demographics (age, sex); primary infectious diagnoses; comorbid conditions; biomarker testing (white blood cell count [WBC], C-reactive protein [CRP], procalcitonin [PCT]); microbiological investigations (culture and sensitivity [C&S] performed, number of cultures, organisms isolated); antimicrobial regimens (empiric vs. targeted, drug names, classes); AMS interventions recommended (dose optimization, de-escalation, discontinuation, diagnostic requests, infection prevention measures); and clinician acceptance status. Data were extracted from handwritten and electronic AMS surveillance forms and entered into a structured database. Raw data were subsequently cleaned and standardized before analysis, including harmonization of antimicrobial names, organism nomenclature, and diagnosis categories.

### Statistical Analysis

Data were analyzed using R statistical programming. Categorical variables were summarized as frequencies and percentages. Between-visit-year comparisons for proportions were assessed using the chi-square test or Fisher’s exact test where expected cell counts were <5. Wilson score 95% confidence intervals were calculated for primary proportions (clinician acceptance rates; meropenem utilization) to account for the smaller 2025 cohort. Continuous variables (age, cultures per patient) were described using median and interquartile range (IQR) and compared with the Mann-Whitney U test (two-sided). A two-sided p-value <0.05 was considered statistically significant. No adjustment for multiple comparisons was applied, consistent with the descriptive surveillance and hypothesis-generating intent of the study.

### Ethics

This study was conducted in accordance with ethical approval obtained from the University of Ghana Medical Centre Institutional Review Board (UGMC/IRBREVIEW/085/25). A waiver of informed consent was granted by the IRB, given the retrospective, de-identified nature of routine surveillance data.

## RESULTS

### Patient Characteristics and Diagnoses

A total of 102 AMS ward round visits were documented in 2024 and 63 in 2025 (Table 1). The 2024 cohort was predominantly female (54.9%), while 2025 visits were predominantly male (61.9%); this shift approached but did not reach conventional statistical significance (p=0.05). Age distributions were similar between years, with a median age of 43 years (IQR 25 –60) in 2024 and 37 years (IQR 30–58) in 2025 (p=0.99). Both cohorts spanned broad age ranges, from infancy to late adulthood (2024: 1–86 years; 2025: 2–89 years), reflecting the work of the UGMC-AMS team.

**Table 1.** Demographic and Clinical Characteristics of ICU Patients Reviewed During AMS Ward Rounds, 2024 vs. 2025.

| Variable | 2024 (n=102 visits) | 2025 (n=63 visits) | p-value |
| --- | --- | --- | --- |
| **Sex, n (%)** |  |  |  |
| **Female** | 56 (54.9%) | 24 (38.1%) | 0.05 |
| **Male** | 46 (45.1%) | 39 (61.9%) | , |
| **Age (years)** |  |  |  |
| **Median (IQR)** | 43 (25–60) | 37 (30–58) | 0.99<sup>a</sup> |
| **Range** | 1–86 | 2–89 | , |
| **Primary Infectious Diagnosis, n (%)** |  |  |  |
| **Sepsis** | 30 (29.4%) | 18 (28.6%) | 1.00 |
| **Pneumonia (all types)** | 32 (31.4%) | 22 (34.9%) | 0.69 |
| **Surgical site infection** | 12 (11.8%) | 0 (0.0%) | 0.004<sup>b</sup> |
| **Ventilator-associated pneumonia** | 0 (0.0%) | 4 (6.3%) | 0.02<sup>b</sup> |
| **Urinary tract infection** | 4 (3.9%) | 2 (3.2%) | 1.00 |
| **Other / Polytrauma / Neurological** | 24 (23.5%) | 17 (27.0%) | 0.69 |

| Comorbid Conditions, n (%) |  |  |  |
| --- | --- | --- | --- |
| Hypertension | 11 (10.8%) | 1 (1.6%) | 0.02<sup>b</sup> |
| Chronic kidney disease | 8 (7.8%) | 2 (3.2%) | 0.34 |
| Diabetes mellitus | 4 (3.9%) | 1 (1.6%) | 0.66 |
| Sickle cell disease | 4 (3.9%) | 0 (0.0%) | 0.16 |
| AMS = antimicrobial stewardship; ICU = intensive care unit; IQR = interquartile range.<br>Pneumonia (all types) includes lobar, bilateral, aspiration, and ARDS-associated pneumonia.<br>Patients may have >1 diagnosis; rows are not mutually exclusive. <sup>a</sup> Mann-Whitney U test. <sup>b</sup> Fisher's exact test. All other p-values by chi-square test. |  |  |  |

Sepsis was the leading primary infectious diagnosis in both years, accounting for 29.4% of visits in 2024 and 28.6% in 2025. Pneumonia of all types (lobar, bilateral, aspiration, and ARDS-associated) was the second most common diagnostic category (31.4% vs 34.9%). Notably, ventilator-associated pneumonia (VAP) emerged as a distinct diagnosis in 2025 (6.3%), reflecting either increased clinical recognition or surveillance capture. The distribution of comorbid conditions differed between cohorts, with hypertension documented in 10.8% of 2024 visits compared with 1.6% in 2025 (p=0.02).

### Diagnostic Stewardship

Biomarker-guided antimicrobial decision-making increased significantly from 86.3% of visits in 2024 to 100% in 2025 (p=0.005), indicating near-universal adoption of inflammatory and acute-phase markers in 2025 (Table 2). WBC was the most consistently obtained marker in both years (70.6% vs 96.8%; p<0.001). CRP utilization increased substantially (7.8% vs 60.3%; p<0.001), as did PCT (2.0% vs 46.0%; p<0.001). Importantly, 2024 CRP and PCT figures are likely underestimates reflecting incomplete documentation in the AMS surveillance form rather than true non-utilization, as discussed in the Limitations section.

**Table 2.**
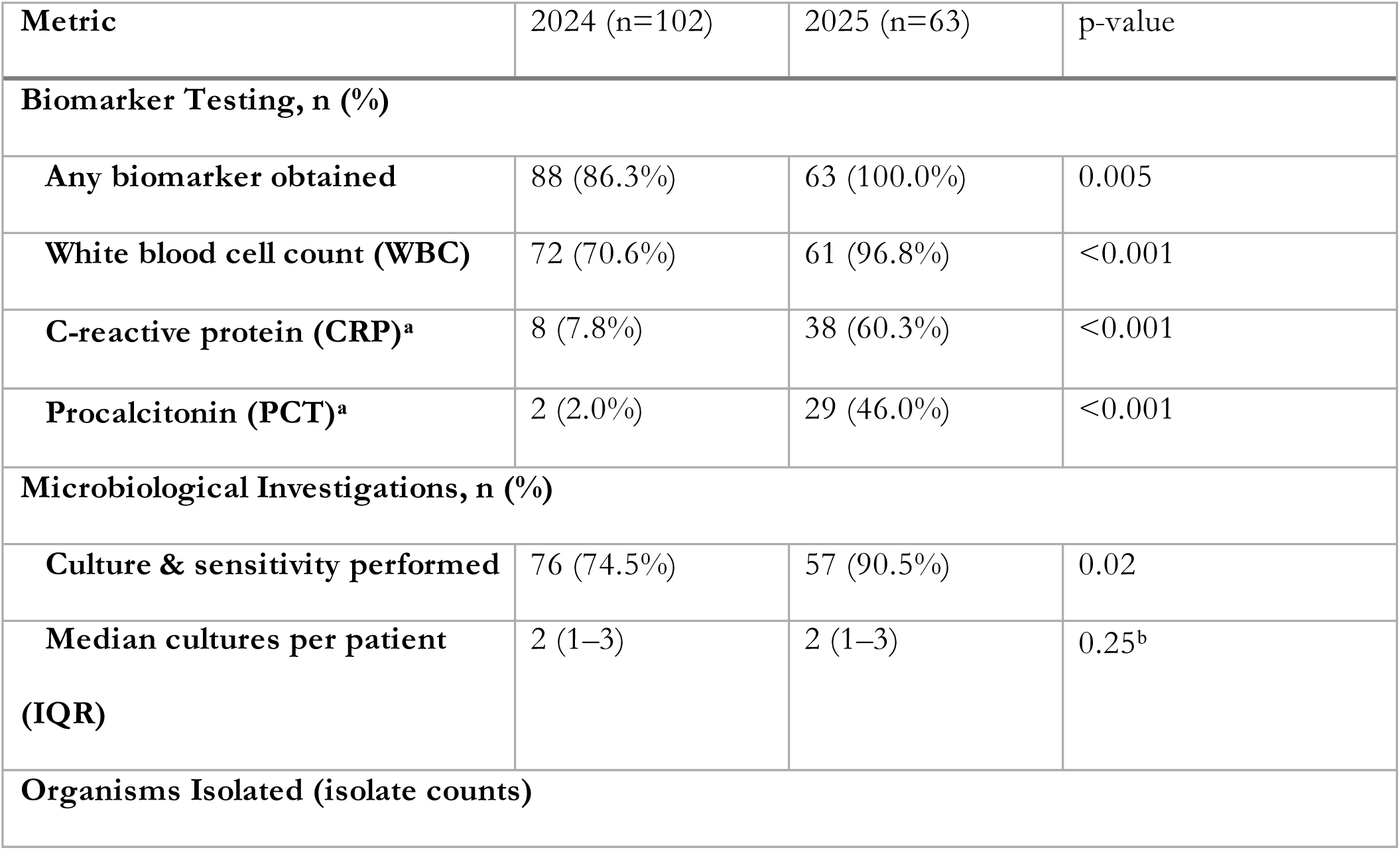
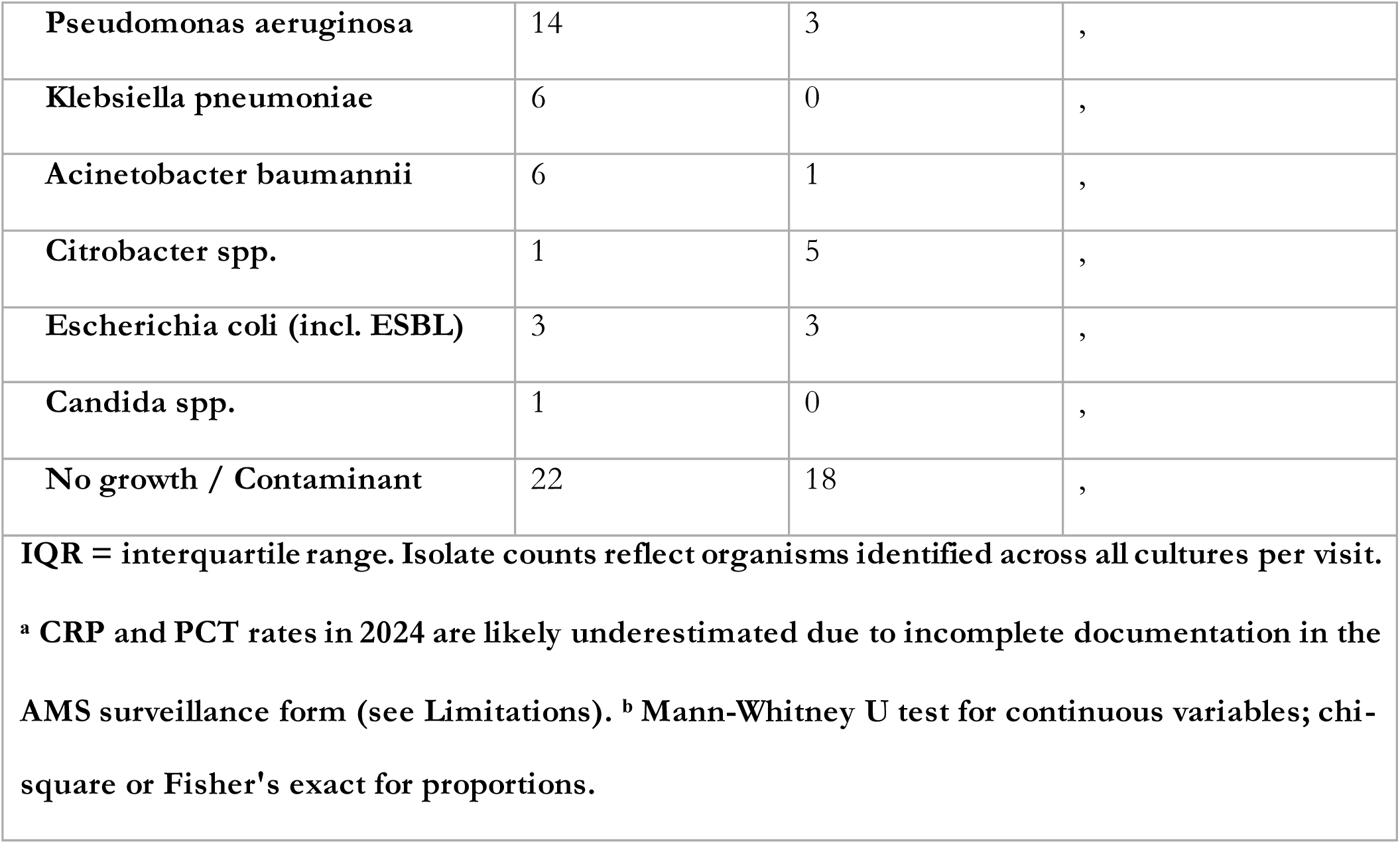
Diagnostic Stewardship Metrics: Biomarker and Microbiological Investigation Utilization.

| Metric | 2024 (n=102) | 2025 (n=63) | p-value |
| --- | --- | --- | --- |
| **Biomarker Testing, n (%)** |  |  |  |
| Any biomarker obtained | 88 (86.3%) | 63 (100.0%) | 0.005 |
| White blood cell count (WBC) | 72 (70.6%) | 61 (96.8%) | <0.001 |
| C-reactive protein (CRP)<sup>a</sup> | 8 (7.8%) | 38 (60.3%) | <0.001 |
| Procalcitonin (PCT)<sup>a</sup> | 2 (2.0%) | 29 (46.0%) | <0.001 |
| **Microbiological Investigations, n (%)** |  |  |  |
| Culture & sensitivity performed | 76 (74.5%) | 57 (90.5%) | 0.02 |
| Median cultures per patient<br>(IQR) | 2 (1–3) | 2 (1–3) | 0.25<sup>b</sup> |
| **Organisms Isolated (isolate counts)** |  |  |  |
| **Pseudomonas aeruginosa** | 14 | 3 | , |
| **Klebsiella pneumoniae** | 6 | 0 | , |
| **Acinetobacter baumannii** | 6 | 1 | , |
| **Citrobacter spp.** | 1 | 5 | , |
| **Escherichia coli (incl. ESBL)** | 3 | 3 | , |
| **Candida spp.** | 1 | 0 | , |
| **No growth / Contaminant** | 22 | 18 | , |
| **IQR = interquartile range. Isolate counts reflect organisms identified across all cultures per visit.**<br>**<sup>a</sup> CRP and PCT rates in 2024 are likely underestimated due to incomplete documentation in the AMS surveillance form (see Limitations). <sup>b</sup> Mann-Whitney U test for continuous variables; chi-square or Fisher's exact for proportions.** |  |  |  |

Culture and sensitivity testing rose from 74.5% to 90.5% of visits (p=0.02), indicating improved pre-antibiotic culture practices. The median number of cultures obtained per visit remained stable at 2 (IQR 1–3) in both years (p=0.25). *Pseudomonas aeruginosa* was the predominant isolated organism in 2024 (14 isolates), with a notable reduction in recovered isolates in 2025 (3 isolates); *Citrobacter* spp. emerged as a more frequent pathogen in 2025 (5 vs 1 isolate). *Klebsiella pneumoniae* and *Acinetobacter baumannii* were isolated predominantly in 2024. No-growth results were common across both years (22 in 2024; 18 in 2025), reflecting diagnostic challenges inherent to critically ill ICU patients receiving prior antimicrobial therapy.

### Antimicrobial Prescribing Patterns

Overall, antibiotic utilization remained persistently high across both years (96.1% in 2024 vs 95.2% in 2025), consistent with the high-acuity patient profile of the unit (Table 3). A significant and clinically meaningful shift in prescribing strategy was observed: the proportion of empiric therapy among antibiotic recipients declined from 77.6% to 58.3% (p=0.02), while targeted (culture-guided) prescribing increased correspondingly from 23.5% to 41.7% (p=0.03). This shift represents a substantial improvement in diagnostic stewardship integration.

**Table 3.** Antimicrobial Prescribing Patterns: Empiric vs. Targeted Therapy and Agent Utilization.

| **Prescribing Metric** | **2024 (n=102)** | **2025 (n=63)** | **p-value** |
| --- | --- | --- | --- |
| **Any antibiotic prescribed, n (%)** | 98 (96.1%) | 60 (95.2%) | 1.00 |
| **Therapy Type (among recipients)** |  |  |  |
| **Empiric** | 76/98 (77.6%) | 35/60 (58.3%) | 0.02 |
| **Targeted (culture-guided)** | 23/98 (23.5%) | 25/60 (41.7%) | 0.03 |
| **Antibiotic Classes / Agents (among recipients), n (%)** |  |  |  |
| **Carbapenems** |  |  |  |
| **Meropenem<sup>a</sup>** | 42/98 (42.9%) | 34/60 (56.7%) | 0.13 |
| **Cephalosporins** |  |  |  |
| **Ceftriaxone** | 27/98 (27.6%) | 9/60 (15.0%) | 0.10 |
| **Penicillins / BLI** |  |  |  |
| **Piperacillin/Tazobactam** | 4/98 (4.1%) | 6/60 (10.0%) | 0.18 |
| **Fluoroquinolones** |  |  |  |
| **Levofloxacin** | 10/98 (10.2%) | 9/60 (15.0%) | 0.52 |
| **Ciprofloxacin** | 6/98 (6.1%) | 5/60 (8.3%) | 0.75 |
| **Nitroimidazoles** |  |  |  |
| **Metronidazole** | 27/98 (27.6%) | 6/60 (10.0%) | 0.02 |
| **Aminoglycosides** |  |  |  |
| **Amikacin** | 2/98 (2.0%) | 8/60 (13.3%) | 0.007 |
| **Anti-MRSA Agents** |  |  |  |
| **Vancomycin / Linezolid** | 3/98 (3.1%) | 6/60 (10.0%) | 0.09 |
| **Polymyxins** |  |  |  |
| **Polymyxin B / Colistin** | 6/98 (6.1%) | 6/60 (10.0%) | 0.38 |
| **Antifungals** |  |  |  |
| **Fluconazole / Amphotericin B** | 1/98 (1.0%) | 5/60 (8.3%) | 0.02 |
| **Median antibiotics per patient (IQR)** | 2 (1–3) | 2 (1–3) | , |
| **BLI = beta-lactamase inhibitor; IQR = interquartile range; MRSA = methicillin-resistant Staphylococcus aureus. Patients may receive >1 agent; percentages are of antibiotic recipients. <sup>a</sup> Wilson 95% CI: 2024: 33.5–52.7%; 2025: 44.1–68.4%.** |  |  |  |

Meropenem utilization increased from 42.9% to 56.7% of antibiotic recipients between 2024 and 2025, though this did not reach statistical significance at the visit level (p=0.13; Wilson 95% CI: 33.5–52.7% vs 44.1–68.4%). The observed trend is clinically notable and consistent with escalating acuity, local resistance pressure from gram-negative MDROs, and expanded targeted use.

Ceftriaxone use declined (27.6% to 15.0%; p=0.10), and metronidazole use fell significantly (27.6% to 10.0%; p=0.02), the latter likely reflecting successful AMS recommendations to discontinue redundant anaerobic coverage. Amikacin use increased significantly (2.0% to 13.3%; p=0.007), consistent with combination therapy against MDRO gram-negative infections. Antifungal prescribing (fluconazole/amphotericin B) also increased (1.0% to 8.3%; p=0.02), reflecting greater culture-directed recognition of *Candida* and other fungal pathogens.

### AMS Interventions and Clinician Adherence

AMS interventions were recommended in 69 visits (67.6%) in 2024 and 40 visits (63.5%) in 2025 (p=0.71; Table 4). The distribution of intervention types was broadly similar between years. De-escalation or discontinuation of antimicrobials was the most common recommendation in both cohorts (40.6% and 47.5% of visits with intervention, respectively). Diagnostic requests, including MRSA screening, blood cultures, and bronchoalveolar lavage, were recommended in approximately one-third of intervention visits in both years. Carbapenem-sparing strategies were recommended in 11.6% and 17.5% of intervention visits in 2024 and 2025, respectively (p=0.37).

**Table 4.** AMS Interventions and Clinician Adherence: Recommendation Types and Acceptance Rates.

| Stewardship Metric | 2024 (n=102) | 2025 (n=63) | p-value |
| --- | --- | --- | --- |
| **AMS intervention recommended, n (%)** | 69 (67.6%) | 40 (63.5%) | 0.71 |
| **Intervention Type, n (% of recommended)** |  |  |  |
| **De-escalation / discontinuation** | 28 (40.6%) | 19 (47.5%) | 0.54 |
| **Dose / frequency optimization** | 18 (26.1%) | 11 (27.5%) | 0.87 |
| **Diagnostic request (culture, MRSA, BAL)** | 22 (31.9%) | 14 (35.0%) | 0.73 |
| **Switch to targeted therapy** | 15 (21.7%) | 9 (22.5%) | 0.93 |
| **Antifungal stewardship** | 6 (8.7%) | 5 (12.5%) | 0.56 |
| **IPC / device care reinforcement** | 11 (15.9%) | 8 (20.0%) | 0.60 |
| **Carbapenem-sparing recommendation** | 8 (11.6%) | 7 (17.5%) | 0.37 |
| **Clinician Acceptance** |  |  |  |
| **Accepted (of recommended), n (%)<sup>a</sup>** | **28/69 (40.6%)** | **27/40 (67.5%)** | **0.01** |
| **Not accepted** | 41/69 (59.4%) | 13/40 (32.5%) | , |
| **Reasons for non-acceptance (documented), n** |  |  |  |
| **Clinical judgement / perceived severity** | 18 | 4 | , |
| **Pending diagnostics / awaiting results** | 12 | 3 | , |
| **Formulary / availability**<br>**constraints** | 5 | 2 | , |
| **Not documented / unclear** | 6 | 4 | , |
| **AMS = antimicrobial stewardship; BAL = bronchoalveolar lavage; IPC = infection prevention and control. Intervention types are not mutually exclusive; percentages may exceed 100%.**<br>**Acceptance denominator = visits with intervention recommended. <sup>a</sup> Wilson 95% CI: 2024: 29.8–52.4%; 2025: 52.0–79.9%. Chi-square test.** |  |  |  |

Clinician acceptance of AMS recommendations improved markedly and significantly between years: from 40.6% (28/69; 95% CI: 29.8–52.4%) in 2024 to 67.5% (27/40; 95% CI: 52.0–79.9%) in 2025 (p=0.01). Non-acceptance was most frequently attributed to clinical judgement or perceived patient severity (18 visits in 2024; 4 in 2025), followed by pending diagnostic results (12 vs 3). Formulary and drug availability constraints accounted for non-acceptance in 5 and 2 visits in 2024 and 2025, respectively.

## DISCUSSION

### Overview of Program Maturation

This two-year comparative analysis of PAF-based AMS surveillance in the UGMC ICU provides longitudinal evidence of a program in active maturation across multiple stewardship domains.

Between 2024 and 2025, biomarker adoption reached universality, culture and sensitivity utilization improved significantly, the proportion of targeted (culture-guided) prescribing nearly doubled, and clinician acceptance of stewardship recommendations rose from 40.6% to 67.5% (p=0.01). Taken together, these findings demonstrate that consistent, multidisciplinary PAF embedded in daily ICU ward rounds can produce meaningful and measurable improvements in prescribing culture and diagnostic practice over a relatively short program lifespan, a finding of relevance given the scarcity of longitudinal AMS data from quaternary LMIC settings.^1,4^

Critically, the improvements observed were not uniform across all domains. Overall antibiotic utilization remained essentially unchanged at approximately 96% in both years, and meropenem prescribing continued to rise. These divergent trajectories, gains in process quality alongside persistence of broad-spectrum use, are not contradictory; they reflect the operational realities of a high-acuity ICU serving a population with limited diagnostic infrastructure, delayed microbiological turnaround, and formulary constraints. Understanding both trajectories is essential to interpreting the program’s progress honestly and planning its next phase.

### Clinician Acceptance: The Central Outcome

The improvement in clinician acceptance from 40.6% (28/69; 95% CI: 29.8 –52.4%) to 67.5% (27/40; 95% CI: 52.0–79.9%) (p=0.01) is the most clinically consequential finding of this study and the most direct indicator of program effectiveness. Acceptance rates in this range are consistent with or exceed those reported from comparable AMS programs in sub-Saharan Africa and other LMIC settings, where first-year acceptance frequently falls below 50% before improving with sustained engagement.^17^ The trajectory observed at UGMC therefore benchmarks favorably against international comparators.

Beyond the aggregate figure, the qualitative shift in the pattern of non-acceptance between years is particularly informative. In 2024, the most frequently cited reason for rejecting an AMS recommendation was clinical judgement or perceived patient severity (18 cases), suggesting that prescribers regarded stewardship recommendations as insufficiently grounded in the clinical context of their patients. By 2025, this category had fallen dramatically to just 4 cases. The dominant residual barriers were instead pending diagnostic results (3 cases) and formulary or drug availability constraints (2 cases), both system-level limitations rather than interpersonal resistance to stewardship. This distinction is clinically and programmatically significant: the AMS team’s recommendations are now broadly regarded as credible, and the outstanding obstacle is ensuring the diagnostic and pharmaceutical conditions exist to act on them.

Multiple mechanisms likely underlie this improvement. Consistent AMS team presence during daily rounds is the single most replicated driver of acceptance in the PAF literature, transforming the stewardship relationship from that of an external auditor to a recognized clinical partner.^12,18^ The compounding effect of iterative feedback, where a clinician who accepted a recommendation in 2024 and observed a positive patient outcome carries that experience into 2025, should not be underestimated. Alignment of recommendations with locally derived microbiological data and the program’s demonstrable track record of culture-guided de-escalation further reduces the grounds for subjective dismissal of stewardship advice.^19^

### Diagnostic Stewardship and the Shift to Targeted Prescribing

The significant increase in culture and sensitivity testing (74.5% to 90.5%; p=0.02) and the near-doubling of targeted (culture-guided) prescribing (23.5% to 41.7%; p=0.03) represent the most structurally important changes in prescribing behavior observed across the two years. These shifts are mechanistically linked: better culture rates provide the microbiological foundation upon which de-escalation and targeted therapy decisions rest. The IDSA/SHEA guidelines identify culture-guided de-escalation at 48–72 hours as a priority strategy specifically because it addresses the principal driver of unnecessary broad-spectrum use, the clinician’s reasonable reluctance to narrow therapy without knowing the pathogen.^7^

The concurrent increase in biomarker-guided prescribing, from 86.3% to 100% of visits (p=0.005), with CRP adoption rising from 7.8% to 60.3% and procalcitonin from 2.0% to 46.0%, provides the complementary inflammatory evidence base that supports antibiotic initiation, continuation, and discontinuation decisions. The markedly lower 2024 rates for CRP and PCT warrant specific comment: these figures likely reflect incomplete documentation in the AMS surveillance form rather than true non-utilization, as discussed in the Limitations section. The 2025 figures, if representative of actual ordering practice, reflect a genuine and meaningful embedding of biomarker-guided clinical reasoning.

The significant reduction in metronidazole prescribing (27.6% to 10.0%; p=0.02) is a concrete and measurable consequence of AMS recommendations to discontinue redundant anaerobic coverage, a common stewardship target when carbapenems already provide adequate anaerobic activity. This reduction represents one of the clearest examples of a stewardship recommendation translating into a sustained prescribing change and validates the operational effectiveness of the PAF model at this institution.^20,21^

### Antimicrobial Prescribing: Persistent Challenges

Despite process improvements, two prescribing patterns require attention. First, overall antibiotic utilization remained essentially unchanged (96.1% vs 95.2%; p=1.00). In isolation, this appears discouraging, but must be contextualized: ICU patients at a quaternary referral center frequently arrive with established infection, hemodynamic compromise, or recent surgical intervention, making antibiotic initiation near-universal at the point of AMS review.^4^ The relevant stewardship objective in this setting is not to reduce clinically appropriate initiation rates, but to optimize the choice, duration, and route of antibiotics already prescribed. The significant shift toward targeted prescribing, the reduction in empiric use, and the decrease in redundant metronidazole co-prescription collectively demonstrate that this optimization objective is being pursued successfully. Second, meropenem utilization increased from 42.9% to 56.7% of antibiotic recipients (p=0.13; 95% CI: 33.5–52.7% vs 44.1–68.4%). While not statistically significant at the visit level, this upward trend is clinically notable and warrants sustained monitoring. The 2025 cohort included emergent cases of ventilator-associated pneumonia and a higher proportion of trauma and neurological cases, all carrying high carbapenem indication rates. The recovery of *Citrobacter* spp., organisms bearing inducible AmpC beta-lactamases conferring third-generation cephalosporin resistance, provides legitimate culture-guided grounds for carbapenem selection in several 2025 cases. ^22^ The concurrent significant increases in amikacin (2.0% to 13.3%; p=0.007) and antifungals (1.0% to 8.3%; p=0.02) further reflect greater integration of culture results into prescribing decisions, a qualitative improvement in prescribing rationale even where the chosen agents are broad-spectrum.

Notwithstanding these contextual factors, the rising carbapenem trend demands a structured response. The AMS team’s carbapenem-sparing recommendations increased from 11.6% to 17.5% of intervention visits (p=0.37), signaling programmatic awareness. Translating this awareness into sustained prescribing change will require formal carbapenem-sparing protocols, rapid diagnostic platforms (multiplex PCR, MALDI-TOF), and structured de-escalation pathways with explicit 48–72-hour review triggers.^23,24^

### Microbiology and Local Resistance Context

The shift in predominant ICU pathogens between years, from *Pseudomonas aeruginosa* (14 isolates in 2024) to a more heterogeneous pattern, including emergent *Citrobacter* spp. (5 isolates in 2025), may reflect genuine shifts in ICU microbial ecology, changes in patient case-mix, or improved culture sensitivity. Regardless of mechanism, this shift carries direct stewardship implications. *Citrobacter* spp., particularly *C. freundii* and *C. koseri*, possess inducible AmpC beta-lactamases, rendering them resistant to third-generation cephalosporins despite in vitro susceptibility at the time of testing, a phenomenon of clinical relevance when ceftriaxone is used empirically. ^25^ The directional decline in ceftriaxone prescribing (27.6% to 15.0%; p=0.10) is consistent with appropriate AMS guidance in the context of this evolving local ecology.

*Pseudomonas aeruginosa* persisted as the leading recovered organism across both years, even with reduced isolate counts, underscoring the need for ongoing anti-pseudomonal surveillance, maintenance of a locally derived antibiogram, and targeted protocols ensuring anti-pseudomonal therapy is promptly de-escalated when cultures confirm an alternative pathogen.

### Lessons Learned: Implications for LMIC AMS Program Implementation

The two-year UGMC experience generates several transferable lessons for institutions implementing or scaling PAF programs in LMIC quaternary ICU settings.

#### Lesson 1: Consistent daily presence is the most important driver of the program

The improvement in clinician acceptance is most plausibly attributed not to any single intervention but to the accumulated relational capital of a team embedded in daily rounds. Institutions beginning a PAF program should prioritize staffing continuity and clinical integration over the comprehensiveness of their surveillance database.

#### Lesson 2: Track the quality of non-acceptance, not just the quantity

The aggregate acceptance rate understates program maturity. A program where non-acceptance is driven by clinical uncertainty is at a different developmental stage than one where non-acceptance is driven by pending laboratory results or drug unavailability. Disaggregating documented reasons for non-acceptance at each surveillance cycle is therefore as important as tracking the rate itself.

#### Lesson 3: Rising carbapenem use in year two should not be interpreted as program failure

In a high-acuity LMIC ICU with evolving MDRO pressure and limited rapid diagnostics, some increase in carbapenem use may be unavoidable and, in culture-guided cases, appropriate. The meaningful metric is not carbapenem use per se, but the proportion of courses that are culture-guided, time-limited, and reviewed at 48–72 hours. Programs should track empiric versus targeted carbapenem days separately.

### Strengths and Limitations

Several limitations must be acknowledged. First, CRP and PCT documentation rates in 2024 are likely substantially underestimated in the surveillance form; cross-referencing with laboratory order records is necessary to establish true utilization rates. Second, the visit-level design precludes patient-level outcome inference, including ICU length of stay, in-hospital mortality, and infection resolution, which remains an important gap to address in future prospective analysis. Third, the smaller 2025 cohort (n=63) reduces statistical power for several comparisons, including the meropenem trend (p=0.13), which may have reached conventional significance in a patient-level or larger dataset.

Fourth, the retrospective single-center design limits causal inference and generalizability. Fifth, no adjustment for multiple comparisons was applied, consistent with the descriptive, hypothesis-generating intent of this surveillance evaluation. Sixth, twelve visits in 2025 lacked a recorded Patient ID, representing a minor but non-zero data quality limitation. Finally, the study did not capture patient-level clinical outcomes, limiting the ability to directly link the prescribing improvements documented here to patient benefit, an essential next step for demonstrating the full value of the program.

## Data Availability

The minimal dataset is available via https://github.com/pmvc23/DATA

## Acknowledgements

The authors thank the nursing and medical staff of the UGMC General Intensive Care Unit for their support in AMS ward round participation and data documentation, and the UGMC Microbiology Laboratory team for their role in culture and sensitivity processing. We are grateful to the patients and families of the UGMC ICU for their implicit trust in the care provided by this program.

## Funding

This study did not receive any specific grant from funding agencies in the public, commercial, or not-for-profit sectors. The antimicrobial stewardship program at the University of Ghana Medical Centre is supported by institutional resources.

## Transparency declarations

### Conflicts of interest

None declared. All authors confirm that they have no financial or personal relationships with other people or organizations that could inappropriately influence this work. Use of artificial intelligence: AI-assisted tools - Grammarly was used in the preparation and editing of this manuscript.

### Author contributions

P.K.M. conceived and designed the study, led data collection, performed the statistical analysis, and drafted the manuscript. N.A.B.O., D.F.O.A., J.M., S.A.D., F.S.B., and G.N.A.O. contributed to data collection, AMS ward round documentation, and critical revision of the manuscript. B.B.Y. contributed to clinical data analysis and critical revision of the manuscript. G.B.K. provided senior oversight, contributed to study design, and critically revised the manuscript for important intellectual content. All authors read and approved the final version of the manuscript.

---

**Figure 1.**
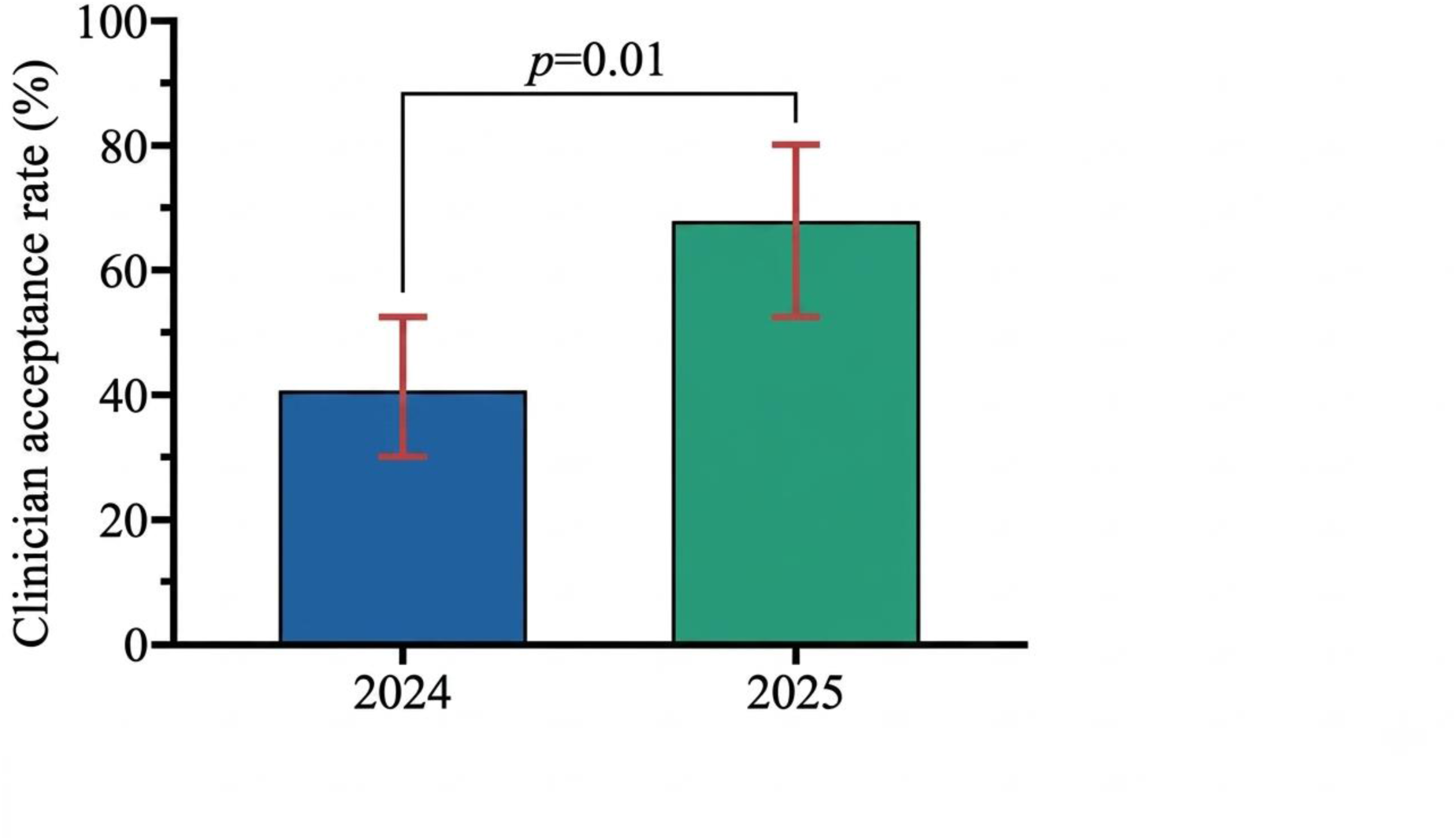
Clinician Acceptance of AMS Interventions, 2024 vs. 2025. Bar chart comparing clinician acceptance rates of antimicrobial stewardship recommendations during prospective audit and feedback rounds in 2024 (40.6%; 95% CI: 29.8–52.4%; n=69 visits with intervention) and 2025 (67.5%; 95% CI: 52.0 –79.9%; n=40 visits with intervention). Error bars represent Wilson score 95% confidence intervals. p=0.01 by chi -square test. AMS = antimicrobial stewardship. Alt text: Bar chart showing clinician acceptance rates rising from 40.6% in 2024 to 67.5% in 2025, with red error bars representing Wilson score 95% confidence intervals and a significance bracket indicating p=0.01.

**Figure 2.**
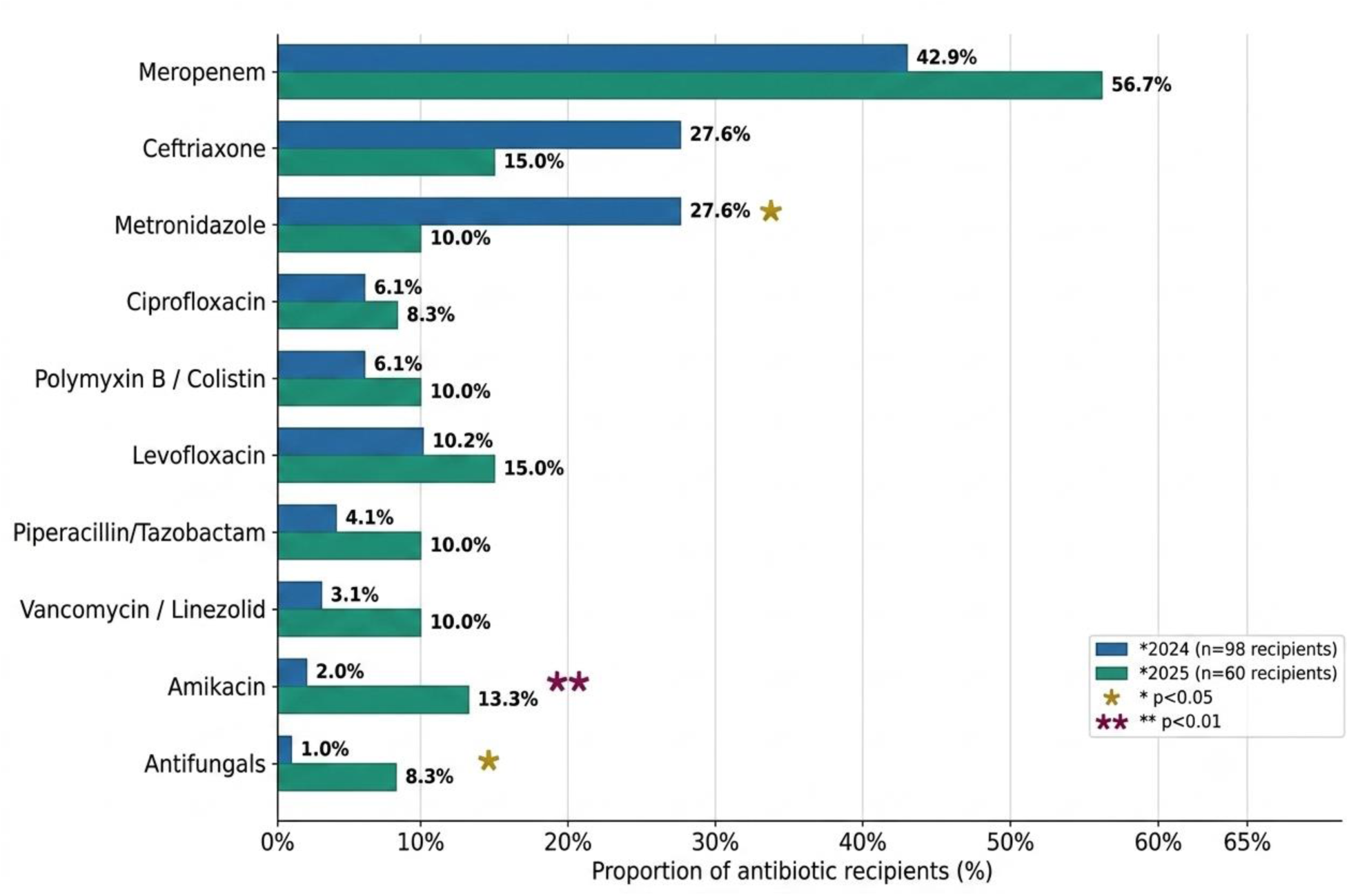
Shift in Antibiotic Class Utilization, 2024 vs. 2025. Grouped horizontal bar chart comparing the percentage of antibiotic recipients prescribed major antibiotic classes in 2024 (n=98 recipients) and 2025 (n=60 recipients). Carbapenem (meropenem) utilization increased from 42.9% to 56.7%; cephalosporin (ceftriaxone) and nitroimidazole (metronidazole) use declined. Statistically significant changes: metronidazole (p=0.02), amikacin (p=0.007), targeted therapy rate (p=0.03). Alt text: Grouped horizontal bar chart comparing antibiotic agent use between 2024 and 2025 among ICU antibiotic recipients. Meropenem use increased from 42.9% to 56.7%; metronidazole decreased significantly from 27.6% to 10.0%; amikacin increased significantly from 2.0% to 13.3%.

**Figure 3.**
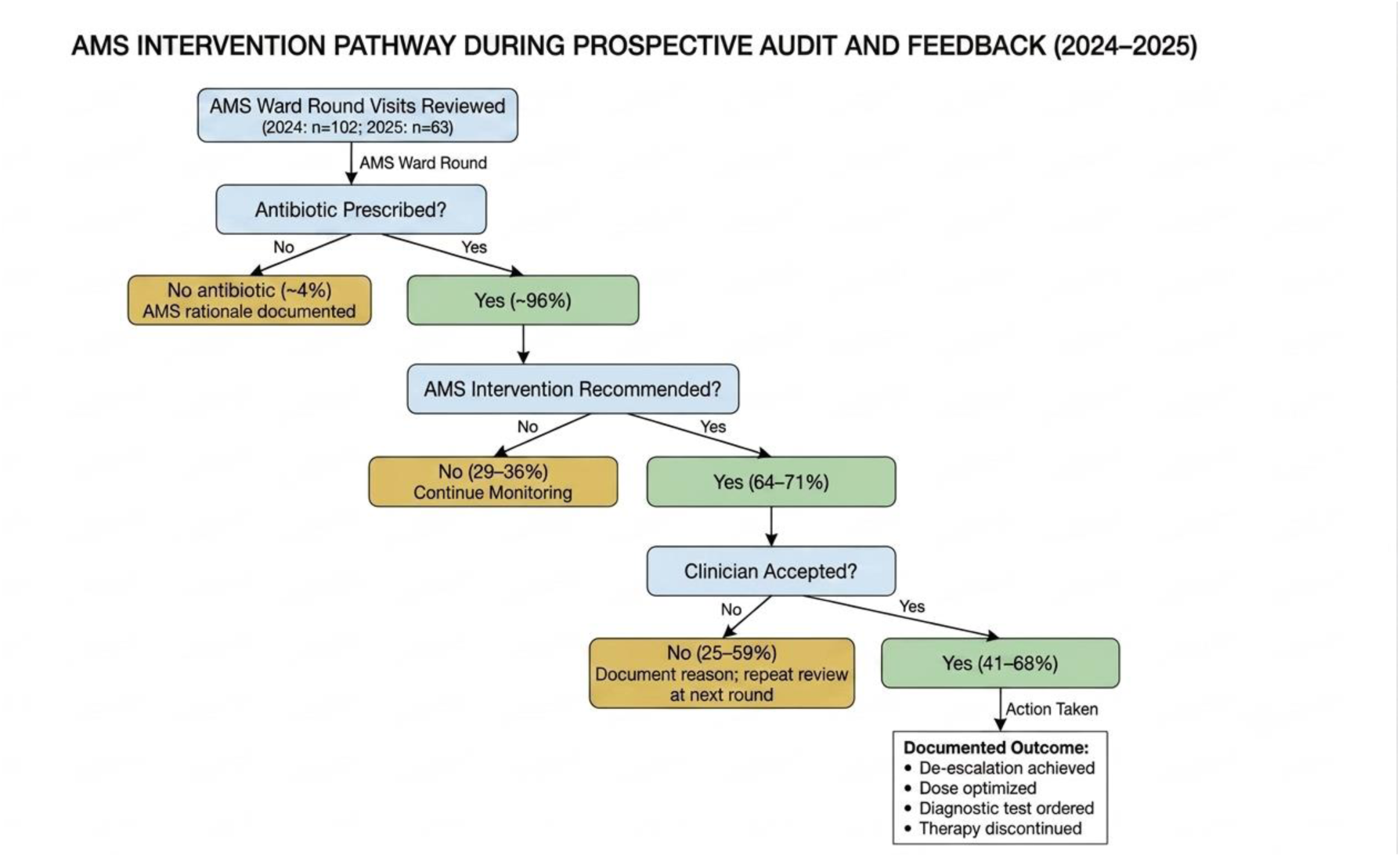
AMS Intervention Pathway During Prospective Audit and Feedback. Flow diagram illustrating the clinical decision pathway for antimicrobial stewardship interventions at the University of Ghana Medical Centre ICU. Nodes represent key decision points; directional arrows indicate the stewardship workflow from patient identification through intervention and outcome documentation. Percentages reflect ranges observed across the 2024–2025 surveillance period. Color coding: blue = decision node; green = accepted intervention outcome; red = non-accepted outcome. AMS = antimicrobial stewardship; BAL = bronchoalveolar lavage; IPC = infection prevention and control; MRSA = methicillin-resistant Staphylococcus aureus. Alt text: Flow diagram of the AMS prospective audit and feedback pathway in the UGMC ICU, showing patient identification, antibiotic prescribing decision, AMS intervention recommendation, and clinician acceptance outcomes across 2024 and 2025.

## Notes

### Competing Interest Statement

The authors have declared no competing interest.

### Author Declarations

This study was conducted in accordance with ethical approval obtained from the University of 137 Ghana Medical Centre Institutional Review Board (UGMC/IRBREVIEW/085/25)

